# Optimizing Lightweight Medical AI for Chest CT Classification: A Distillation and Quantization Approach

**DOI:** 10.1101/2025.11.23.25340844

**Authors:** Isha Kansal, Vikas Khullar, Gifty Gupta, Deepali Gupta, Sapna Juneja, Aimin Li, Saurav Mallik

## Abstract

Medical imaging has been crucial in the diagnostics of pulmonary diseases and the use of chest CT scans is a fundamental diagnostic tool in lung cancer and COVID-19. The clinical importance of the deep learning models used to classify CT images is still hard to deploy because of the high-computational requirements and overfitting. The latest state-of-the-art CNN models, including DenseNet121 and NasNetMobile, use the train model with near-perfect accuracy yet have poor generalization and demand large memory footprint (>2.4 GB), making them unfeasible to apply in healthcare settings with limited resources. To solve this issue, we introduce an end-to-end knowledge distillation and post-training quantization system that can convert the large, overtrained electronics teacher models into small and generalized student networks that can be deployed to the real world to accomplish medical AI. Knowledge distillation allows the student models to study hard labels, as well as softened probabilistic outputs of the teacher, enhancing generalization and reducing overfitting. The post-training quantization also minimizes the model size by shrinking both weights and activations to the 8-bit precision to allow inference with low accuracy degradation. The experiments were run on the Chest CT-Scan Images Dataset (1,252 samples, balanced classes of COVID-19 and non-COVID-19) of the Kaggle standardized and augmented to evaluate well. A variety of teachers were trained (DenseNet121, ResNet50, EfficientNetB3, VGG16/19, Xception, and NasNetMobile) and distilled into small students and quantized to be deployed. The presented pipeline reduced the memory usage (approximately 2,465 MB to approximately 618 MB) by a factor of 4 with the quantized DenseNet121 student being able to reach 91.4% validation accuracy as compared to its teacher (77.2%). There was also better generalization by distilled students, where EfficientNetB3 and NasNetMobile attained +42% and +30% validation gains respectively. This paper offers a deployable and resource-efficient medical AI architecture, which is capable of striking a balance between diagnostic accuracy and computational efficiency. The findings reveal that knowledge distillation and quantization can be combined to provide lightweight, high-performing chest CT classifiers to mobile CT devices, edge devices, and low-resource clinical settings as one step towards closing the gap between research-level AI systems and those that can be deployed significantly more effectively in the clinic.

## 1. Introduction

Medical imaging has also come out to be one of the most effective diagnostic tools in the health sector to enable clinicians to view the internal structures of the human body in an extraordinarily detailed way. Amongst the other imaging modalities such as X-ray, Magnetic Resonance Imaging (MRI) and ultrasound, the chest Computed Tomography (CT) has remained an indispensable aid to the diagnosis of pulmonary conditions. It is established that CT technique is a high-resolution three-dimensional imaging of the thoracic cavity that is specifically useful in the diagnosis of such diseases as lung cancer, tuberculosis, pulmonary fibrosis, Chronic Obstructive Pulmonary Disease (COPD), and infectious diseases, including COVID-19 [1]. The CT of the chest provides a clear information on internal structure of the lungs and plays central role in diagnosis of various diseases of lungs. These scans show minimal changes in tissue density, lesion shape, and patterns of lung involvement which are essential in the proper diagnosis and treatment planning. The examples presented in Figure 1 are some of the typical cases in various categories that are found in clinical practice. The initial group of images (a-c) depicts the normal lung tissue, which serves as a control image of a healthy anatomy. The second group (d-f) presents the cases of adenocarcinoma or one of the most frequent types of lung cancer in which the abnormal growths and irregular appearances of the tissue structures are clearly exposed. The third group (g–i) shows large cell carcinoma, which include cells that are larger and poorly differentiated and that are usually difficult to diagnose. The last set (j l) contains other variants or less typical appearances, the concept of heterogeneity of the lung pathology. The visual summary of these classes by the figure indicates that the CT scan of the chest is complex and variated, which is why automated and accurate classification techniques are required. This heterogeneity is the basis of the AI-based methods reviewed in this paper where powerful models need to be trained to identify and distinguish such diverse patterns that can support healthcare experts in making well-informed clinical judgments.

**Figure 1:**
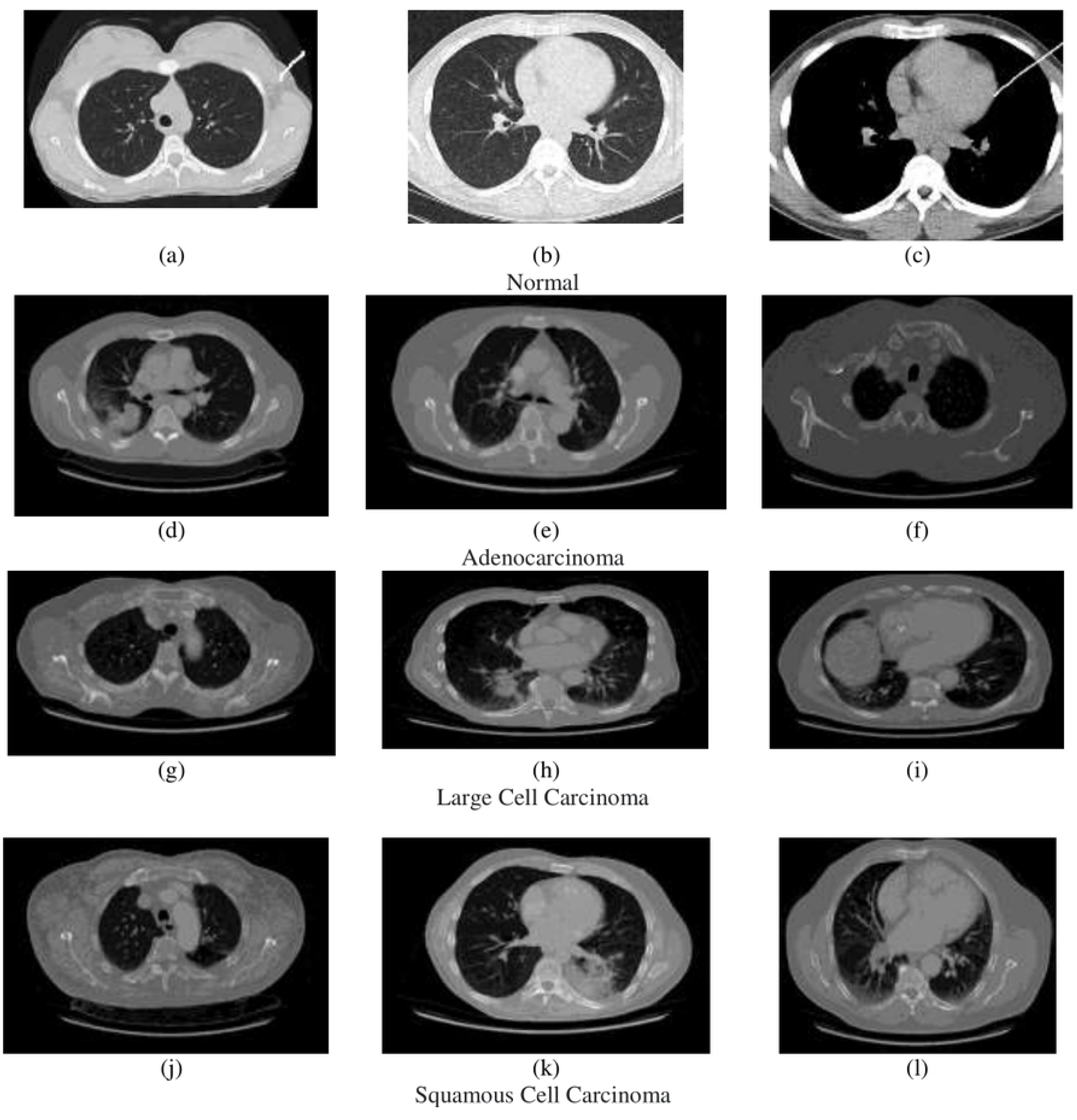
Representative chest CT scan images across different categories. The images (a-c) represent the normal lung tissue that is used as a reference in healthy anatomy. Images (d f) depict the cases of adenocarcinoma, with the emphasis on abnormal formations and irregular pattern of lungs. Large cell carcinoma (g-i) is a tumor with poorly differentiated cell structures that are represented in the images. Images (j -l) reflect other forms of pathological variations or other conditions that have a common occurrence in the clinical practice.

During the past few years, explosive use of Artificial Intelligence (AI) in medical imaging has been among the most effective and accurate Computer-Aided Diagnosis (CAD) systems. CNNs and deep learning, in particular, have proven to have extraordinary powers to automatically learn discriminative components in medical pictures, which is better than feature-based techniques developed by hand [2]. More successful architectures in terms of medical image classification, segmentation and detection have been demonstrated to be VGG, ResNet, DenseNet, EfficientNet and Xception. These models can be used to model complicated trends in imaging data that might be hidden to human specialists, and that is why the models assist radiologists to reduce diagnostic mistakes and improve patient outcomes [3]. With the growing access to CT scans of the chest, deep learning-based classification systems possess a vast potential in the field of early detection and massive screening of the lung-related diseases. Nevertheless, even though AI has a remarkable potential in healthcare, there is still a considerable gap between the research-quality AI models and the implementation of clinical solutions. Such a gap mainly comes because of the computational and structural requirements of contemporary deep learning architectures [4]. State-of-the-art CNN architectures have demonstrated impressive performance in benchmarks datasets but with poor generalization ability and high resource usage, are not suitable to be deployed in real medical practice. To begin with, a significant number of large-scale models tend to overfit. To illustrate the point, in our experiments the DenseNet121 teacher model has 100 percent training accuracy and only 77.2 percent validation accuracy, and this means that the model was memorizing the training examples instead of learning generalizable features. Such a pattern was replicated with NasNetMobile that had accuracy of 99.7% in training, whereas it had accuracy of 63.5% in validation. The issue of overfitting is especially important in clinical environments when a model has to be generalizable to the unknown data of various populations, imaging procedures, and equipment. Second, large deep models have significant computational cost. VGG19 and DenseNet121 models use more than 2.4 GB of memory at inference [5]. Such models require either a high-end GPUs or special accelerators that are frequently not available in healthcare facilities with limited resources. Scalability is limited since even in well-equipped hospitals, the use of costly hardware limits scalability, especially when it is necessary to use models to process high volumes of CT scans in real time. Third, these challenges are even worsened by deployment constraints. Medical devices, peripherals, and medical clinics in rural settings commonly have a small amount of processing capacity. Indicatively, mobile CT scan vans used in remote areas cannot accommodate the large-sized GPU-based systems. On the same note, real-time AI is required in latency-sensitive applications, like emergency triage to deliver results in seconds. In this regard, conventional heavy CNNs are unsuitable in clinical translation. These shortcomings confirm the necessity of lightweight AI systems that balance predictive accuracy with computational performance [6]. It is very important to work on lightweight models of medical imaging to narrow the divide between the development of research on medical imaging and its actual use. The main notion is to maintain diagnostic accuracy and simultaneously lower computational and memory demands to the extent, which is applicable to the real-world. In this quest, two complementary methods have emerged to the forefront: knowledge distillation and model quantization [7] [3].

Knowledge distillation was proposed by Hinton et al. [8], and it is a type of training where the small student model is trained to behave like the large teacher model. The student model does not learn directly by the ground truth labels only but also takes advantage of the so-called soft labels or probabilistic outputs of the teacher, which conveys more information about the relations between the classes. Such imparting of knowledge allows the student to perform at the performance levels that are almost equal or even greater than its teacher despite having fewer parameters and lower complexities. With respect to medical imaging, distillation can produce models that generalize more efficiently, through eliminating overfitting, which is an important feature of clinical deployment. On the other hand, the model quantization is a post training optimization method that downsizes the model size by lowering the numerical accuracy of model weights and activations, commonly 32-bit floating-point numbers to 8-bit integers. This significantly decreases the memory footprint and speed in inference, at times with only a small decrease in accuracy. As an example, quantizing student models in our experiments saved memory resources by almost 75 percent (∼2465 MB to ∼618 MB) and still retained accuracy of within 1-3 percent of the original. These cuts are essential in deploying models on embedded systems, edge devices and low-power clinical platforms. Use of a combination of distillation and quantization yields a powerful framework of developing models which are accurate and deployable. The twofold attitude is in correlation with the long-term aim of democratizing medical AI by ensuring the availability of leading-edge technologies in a wide range of healthcare facilities, such as those that have scanty computational resources. A systematic implementation has been offered in this work to create deployable AI models to classify chest CTs using knowledge distillation and post-training quantization [3]. The novelty of our work are outlined below:

- **Teacher–Student Knowledge Distillation Framework:** We trained multiple teacher networks—DenseNet121, ResNet50, EfficientNetB3, VGG16, VGG19, Xception, and NasNetMobile—and distilled their knowledge into student models. The select group of students was greatly superior to their teachers in terms of validation performance. As an example, DenseNet121 went up to 90% (student) against 77.2 (teacher), where EfficientNetB3 went up to 92 against 49.2.
- **Improved Generalization:** The student models had strong performance compared to the teacher models that were most of the time overfitted. As an example, NasNetMobile improved its validation accuracy (teacher) of 63.5% to 93% (student) with a difference of 29, which illustrates the enhanced generalization ability.
- **Memory efficiency:** Post-Training Quantization of the computation results makes the computation results quantized, thereby saving memory space.
- **Memory:** A memory-saving technique is to post-quantize the results of the computation, and thus the computation results are quantized. The student models shrunk in memory size by almost three-quarters, due to quantization, as thepreviously 2465-MB models became only 618-MB. Although there was this compression, there was minimal performance degradation. An example of quantized DenseNet121 achieved a validation accuracy of 91.4 percent, an improvement of 1 percent over 90 percent before quantization. The overall performance of the business can be assessed by examining all its activities and evaluating the success of each segment.
- **Extensive Performance Analysis:** Our comparative analysis was done in strict terms of teacher, student, and quantized models in relation to the most important performance measures, which included accuracy, precision, recall, training and validation losses, and memory footprint. This discussion gives a comprehensive insight of the trade-offs that exist between performance and efficiency in the design of deployable medical AI systems. The article discusses deployable AI in healthcare and how it has evolved in the past few years.
- **Deployable AI in Healthcare:** Our pipeline is lightweight, resource-efficient, and has high diagnostic accuracy by combining distillation and quantization. These models are applicable to workflows in clinical departments, mobile imaging devices, and edge-computing gadgets, and they bring AI-driven diagnostics one step nearer to the application in the real world of various healthcare settings.

In sum, this paper is a step in the direction of deployable medical AI as it shows that advanced compression methods can be used to convert big, overfitted models into smaller, generalized, and efficient ones. Through knowledge distillation and quantization in the classification of chest CT, we point out the way to construct AI solutions beyond the scope of accuracy but also scalable and available. The work we have conducted fits into the emerging aspect of resource-efficient medical AI, opening the door to the broader use of AI in clinical settings and in environments with limited resources and at the edge.

## 2. Related Work

Deep learning has proven to be extremely successful in the analysis of CTs of the chest, and CNNs and more recent architectures like DenseNet, ResNet, and EfficientNet have already demonstrated impressive performance in the lung disease diagnosis, including cancer and COVID-19. Nonetheless, these models are computationally expensive, they consume a large amount of memory and are also susceptible to overfitting, limiting their application to the edge devices or in low-resource environments. Responding to this, compression methods of models have been looked into: knowledge distillation (KD) is a method of transferring knowledge between large (teacher) networks to lightweight (student) networks to enhance generalization and efficiency, whereas quantization saves memory and computation with representations in lower precision. Even though KD and quantization have been used independently in medical imaging, there has been little systematic integration of both in the classification of chest CT. The previous work mainly focuses on accuracy, and neglects deployment-important measures, including memory footprint, inference latency and robustness. This introduces a tradeoff where efficient pipelines that are a combination of KD and quantization are required to trade off accuracy against resource efficiency to be able to scale and deploy AI to real-world medical imaging needs. Table 1 highlights some of the methodologies, data sets, model structures, performance indicators and deployment factors found in the literature. It raises the merits and shortcomings of the previous research, such as the challenge of overfitting, limited memory, and the inability to combine accuracy and deployability. This review paper puts it in perspective and purpose behind the suggested knowledge distillation and quantization framework that could be used to mitigate significant gaps in clinical AI application to lung disease diagnosis.

**Table 1:** Summary of recent studies on chest CT and related medical imaging classification.

| Reference No. | Methodology/Model | Dataset/Population | Key Findings/Contributions | Remarks |
| --- | --- | --- | --- | --- |
| [9] | Multi-classification deep learning models (VGG19+CNN, ResNet152V2, ResNet152V2+GRU, ResNet152V2+Bi-GRU) | Public chest X-ray and CT datasets with 4 classes (Normal, COVID-19, Pneumonia, Lung Cancer) | Proposed first multi-class model distinguishing COVID-19, Pneumonia, and Lung Cancer. VGG19+CNN achieved best results: 98.05% accuracy, 99.66% AUC. | Demonstrated benefit of combining X-ray and CT images; improved early detection. |
| [10] | Multi-view Knowledge-based Collaborative (MV-KBC) deep model using ResNet-50 submodels | LIDC-IDRI benchmark dataset (lung nodule CT scans) | Decomposed 3D lung nodules into 9 views, used adaptive weighting of submodels. Achieved 91.6% accuracy, 95.7% AUC, outperforming 5 SOTA methods. | Tackled limited data issue via knowledge-based collaborative learning; improved false negative rate reduction. |
| [11] | Deep Learning Chest Disease Detection Network (DCDD_Net) with Borderline-SMOTE for imbalance | 20 publicly available datasets (X-ray, CT, cough sound) | Identified 9 diseases (COVID-19, LC, Pneumonia, TB, etc.). DCDD_Net achieved 96.67% accuracy, 95.61% F1, 99.43% AUC; outperformed EfficientNet-B0, DenseNet-201, InceptionResNet-V2, Xception. | Novel integration of cough sound scalograms with X-ray and CT; robust via statistical validation. |
| [12] | Knowledge Distillation (KD) with CNNs & Transformers, Explainable AI (XAI) heatmaps | ChestX-ray14, CheXpert, PadChest datasets | Proposed lightweight student model trained via KD. Using DenseNet161 teacher, EEEA-Net-C2 achieved AUC of 83.7% (ChestX-ray14), 87.1% (CheXpert), 88.7% (PadChest) with only 4.7M params. | Enabled real-time on-device chest X-ray classification with explainability (heatmaps). |
| [13] | Knowledge Distillation (KD) with multi-task architectures and saliency maps | ChestX-ray14 dataset (14 weakly annotated thoracic diseases) | Compared baseline, standard KD, reversed KD, and self-training KD. Self-training KD improved AUC by up to 6.39% over baseline. Provided saliency maps for interpretability. | First to apply KD for multi-class thoracic abnormality classification with explainability. |
| [14] | VGG-16, DenseNet-161, ResNet-18 models for classification and severity prediction | 2271 chest X-ray images (Normal, Pneumonia, TB, COVID-19) from Clinico Diagnostic Lab, Mumbai, verified by radiologists | Achieved 95.9% (VGG-16), 98.9% (DenseNet-161), and 76% (ResNet-18 for severity). Enabled multi-class COVID vs other diseases classification and severity-level detection. | Demonstrated potential of X-rays for low-cost, rapid COVID-19 screening. |
| [15] | U-Net for lung segmentation + ResNet-50 classification with custom rules | 222 patients (116 COVID-19, 88 CAP, 127 Normal); ~70k CT slices from IEEE ICASSP Challenge | Accuracy 93.84% on test set. Performed better than novice radiologists and nearly equal to experts. Faster diagnosis (35 min vs 75–93 min). | Strong AI decision support tool for radiologists in CT-based COVID-19 pneumonia detection. |
| [6] | Semantic Similarity Graph Embedding (SSGE) with CNN + Graph Convolutional Network under Teacher–Student mechanism | Benchmarks: ChestXray-14, CheXpert datasets | Explored semantic similarity across images for multi-label chest X-ray classification. Outperformed prior state-of-the-art baselines. | First attempt at leveraging multi-image semantic similarity for better diagnosis consistency. |
| [16] | Pipeline: U-Net segmentation → ResNet-50 classification → GradCam localization → Unsupervised clustering | Development: 50 CT scans (China), Test: 109 COVID + 90 non-COVID patients (Zhejiang); 6150 slices for lung segmentation training | Detected, localized, and quantified COVID-19 lesions. Introduced “Corona score” for severity. Weakly supervised method effective with limited data. | Supports radiologists in disease quantification and severity analysis using CT scans. |
| [17] | Ensemble Deep Learning (EfficientNet-B0, ResNet50, DenseNet121 with attention) | Public CT datasets (LIDC-IDRI, LUNA16) | Proposed ensemble for lung nodule classification; achieved >91% accuracy, AUC >94%. Outperformed single models. | Addressed small nodule detection challenges. |
| [18] | Hybrid CNN-RNN with attention | Private hospital CT dataset + public LIDC-IDRI | Integrated CNN feature extraction with RNN temporal context; accuracy 89.7%, AUC 92.3%. | Showed improved handling of sequential CT slice data. |
| [19] | Multi-task deep learning (segmentation + classification) | LIDC-IDRI dataset | Simultaneous segmentation & classification of nodules; achieved accuracy 90.5%, DSC 0.87. | Reduced false positives; clinically relevant workflow. |
| [20] | Collaborative Deep Learning (CDL) with ResNet-50 submodels, logistic regression for shape features | Chest CT nodules (limited dataset) | CDL achieved 93.24% accuracy in classifying malignant vs benign nodules, outperforming SOTA methods. | Adaptive weighting of submodels reduced false positives/negatives ; feasible for clinical workflow. |
| [21] | Combined AI model (3D CNN features + radiomics + clinical data, stacked ensemble with AutoGluon) | Three datasets: LUNA16 (1004 nodules), LNOP (1027 nodules), LNHE (1525 nodules) | Benign vs malignant classification: 92.8% accuracy; Pathological subtype classification: F1 = 75.5%; Lung-RADS classification: F1 = 80.4%. | First study integrating DL, radiomics, and clinical data; robust across multiple tasks. |
| [22] | Stacked ensemble meta-classifier with EfficientNet feature extraction + kernel PCA + feature fusion. Two-stage approach: RF + SVM → Logistic Regression | Large publicly available CT and CXR datasets (COVID vs Non-COVID) | Achieved superior generalization compared to single CNN models; robust performance across CT and X-ray. Suitable for point-of-care use. | Tackled the problem of small datasets and weak generalization in prior work. |
| [23] | Transfer learning using pre-trained CNN models (ResNet, VGG, Inception, etc.) + augmentation | Kaggle COVID-19 radiography dataset (13,975 images: COVID-19, Viral Pneumonia, Lung Opacity, Normal) | Proposed lightweight CNN variants with good accuracy (up to 96% for multi-class). Demonstrated better performance with fewer parameters. | Balances accuracy and computational efficiency for resource-limited hospitals. |
| [24] | DenseNet-121 with weighted loss + global average pooling, trained with Adam optimizer | CheXpert dataset (224,316 chest radiographs, 65,240 patients); tested on pulmonary nodules and cardiomegaly | Outperformed baseline methods in detecting nodules and cardiomegaly. Improved AUC despite class imbalance. | Focused on misdiagnosis reduction; useful in Valley Fever differential diagnosis. |
| [25] | Intensity map projection (InceptionResNetV2) , Multi-task multi-decoder U-Net segmentation, Prognosis models (XGBoost, Random Forest, Extra Trees, Logistic Regression) | CC-CCII dataset (2,471 patients, 514,103 CT slices), Segmentation subset (1302 scans), Prognosis subset (136 COVID-19 patients), Stony Brook University dataset (241 COVID-19 patients) | Diagnosis accuracy 96%; Prognosis AUC improved to 0.80 (CC-CCII) and 0.81 (SBU) when combining demographics + CT features + lesion segmentation; Fibrosis extent most predictive | CT features with demographics improve prognosis prediction; small deceased patient cohort in CC-CCII is a limitation |
| [26] | Knowledge distillation with ensembles of CNNs (U-Net, ResNet, Dilated CNNs; diverse & uniform ensembles distilled to single CNN) | Chest CT (4 organs), Brain MRI (6 structures), Cardiac cine-MRI (3 structures) | Distilled CNNs maintained ensemble performance while being smaller & faster; both diverse and uniform ensembles outperformed single CNNs; knowledge distillation improved efficiency | Demonstrated feasibility of compressing ensembles for medical segmentation tasks |
| [27] | Transformer-based architectures (ViT, DeiT, BeiT) with distillation techniques | Brain MRI dataset (3264 images, 4 classes: glioma, meningioma, pituitary, no tumor) | Distillation improved accuracy of transformers on small datasets (DeiT +2.2%, BeiT +1% vs ViT baseline); better detection of non-patient cases; showed robustness in limited data settings | Highlights benefit of distillation tokens in improving transformer classification in medical imaging |
| [28] | COVLIAS 3.0: Cloud-based quantized hybrid UNet3+ (VGG-UNet3+, ResNet-UNet3+) | 3,500 CT scans (training), 500 unseen CT scans (testing) from COVID-19 patients | ResNet-UNet3+ improved Dice and BCE loss by 17% and 10% over UNet3+; quantization reduced model size (36–66%); robust via multiple statistical tests ($p < 0.001$) | Cloud-deployable, quantized hybrid models enhanced COVID-19 lesion detection in CT |
| [29] | Predictive tree-structured vector quantization (PTSVQ) for lossy compression of CT scans | CT chest scans (diagnostic evaluation by radiologists) | Images compressed from 12 bpp to 1–2 bpp without significant diagnostic accuracy loss; diagnostic accuracy measured via radiologist performance (nodules, adenopathy) | Early foundational study (1993) on compression impact in medical imaging |
| [30] | Survey of Self-explainable AI (S-XAI) for medical image analysis (input, model, and output explainability via feature engineering, knowledge graphs, attention, concept- and prototype-based learning) | Review of 200+ studies across modalities (X-ray, CT, MRI, Ultrasound, Dermatology, etc.) | First comprehensive survey of S-XAI methods; introduced taxonomy (input/model/output explainability); highlighted applications (lesion segmentation, diagnosis, report generation, VQA); identified challenges & future directions | Provides broad state-of-the-art overview and framework for S-XAI in medical imaging |
| [31] | Dataset Distillation methods (Dataset Condensation - DC, Matching Training Trajectories - MTT); ConvNet architecture | MedMNIST datasets (PathMNIST, DermaMNIST, OCTMNIST, BloodMNIST, TissueMNIST, OrganAMNIST, OrganCMNIST, OrganSMNIST); multimodal medical imaging data | Demonstrated that dataset distillation can significantly reduce dataset size while maintaining comparable performance to full datasets; DC outperformed MTT in many cases; strong correlation found between random selection performance and distillation success; potential for efficient and secure medical data sharing | Dataset distillation is promising for medical imaging, but effectiveness varies by dataset (better on large inter-class variation datasets, less effective on small variation ones); highlights need for tailored approaches |
| [32] | Lightweight CNN with quantization & pruning for portable liver fibrosis diagnosis | Ultrasound elastography images (200 patients) | Model reduced size by 68% with <3% accuracy drop; suitable for deployment on edge devices | Highlights potential for resource-efficient liver fibrosis diagnosis in low-resource settings |
| [43] | Chest CT — lung nodule detection & classification | LIDC dataset | Enhanced Region-based Fully Convolutional Network (RFCN) backbone + a novel multi-layer fusion Region Proposal Network (mLRPN), with median-intensity projection and deconvolutional layers to select candidate ROIs | High performance on LIDC; strong detection + classification; heavy model, may be resource-intensive for deployment |
| [44] | Multi-modal imaging (PET-CT, CT) — image retrieval & visualization (non-classification) | Non-small cell lung cancer dataset for PET-CT/CT retrieval | Content-based image retrieval (CBIR) with 2D + proposed 3D volume visualization; rendering parameters optimized to highlight structures deemed “important” in retrieval process | Focused on visualization & retrieval rather than classification per se; contributes to user interpretability and use in diagnostic support tools; helps reduce cognitive load for users comparing volumes |
| [45] | MRI — brain tumor segmentation and 3D | 496 MRI scans from Bab El Oued University | Preprocessing (Gaussian filtering), Active Geometric Contour Models + morphological operations for segmentation; then 3D reconstruction with 3D slicer and AR display of tumor classes | Emphasis on AR + visualization, not CT chest classification; high-accuracy segmentation and effective 3D visualization; useful reference for improving interpretability or user-interaction in medical imaging systems |

The literature survey shows that there are a few gaps that are critical, which are systematically addressed in the piece of work. Although there are many studies that have shown that deep learning models can be successfully applied to classify chest CTs with a range of accuracies of up to 89-99, many of them are more oriented towards optimal diagnostic applications than a placement of deployment restrictions. Among notable exceptions, there are LS4 and LS5 to discover the use of knowledge distillation in the analysis of chest X-rays, and LS26 to use quantization in the segmentation of COVID-19 CT volumes. Nonetheless, they are either works about X-ray modalities or they are about segmentation tasks but not classification. Moreover, although multiple studies (LS24, LS25) have examined the knowledge distillation within medical imaging, none of them has explicitly used distillation and quantization together to target the specific case of CT chest classification in order to generate deployable solutions. Overfitting of large models is also a worrying tendency identified in the literature with the wide range of discrepancies in the training and validation accuracy being reported in various studies. Our contribution fills this gap by introducing the first attempt at systematic knowledge distillation and post-training quantization in chest CT classification to explain how to shift overfitted teacher models to generalizable and lightweight student networks that can be applied in practice in a clinical setting and retain diagnostic accuracy. The literature review indicates that the use of clinical AI in the classification of chest CT has serious gaps that restrict its implementation. Large networks such as DenseNet and ResNet are the most accurate but their large memory requirements (>2.4 GB) are unfeasible in resource-limited systems. Chest CT Knowledge distillation has not yet been explored in depth, much of the research has been done in X-ray modalities and combination methods combining distillation with quantization have not been tried. Most critically, existing research prioritizes accuracy over deployment feasibility, lacking comprehensive analyses of the accuracy-memory trade-offs essential for real-world medical AI systems.

Knowledge distillation in medical imaging remains underexplored, with only selective applications demonstrated in the literature. LS4 successfully applied distillation to chest X-ray classification, achieving competitive performance with the lightweight EEEA-Net-C2 student model (4.7M parameters) compared to larger DenseNet161 teachers. LS5 extended this to multi-task architectures, showing up to 6.39% AUC improvement through self-training distillation while providing interpretability via saliency maps. LS24 demonstrated ensemble distillation across multiple medical modalities (chest CT, brain MRI, cardiac cine-MRI), proving that distilled models could maintain ensemble performance with improved efficiency. LS25 explored transformer-based distillation for brain MRI, achieving +2.2% accuracy improvement with DeiT over ViT baseline on small datasets.

Medical imaging deployment has a lot of potential in quantization. LS26 applied quantized hybrid UNet3+ models that reduced Neighboring sizes with 36-66% with no difference in accuracy (p<0.001) when compared to COVID-19 CT segmentation. LS27 showed a compressible CT scan between 12 bpp and 1-2 bpp with no loss of diagnostic accuracy whereas LS23 had a 68% model size reduction with less than 3% loss of accuracy in diagnosing liver fibrosis, which makes models approposite to edge devices. The methods of pruning are not as widely researched in the reviewed literature, with LS23 as the main example of pruning with quantization as a method of portable medical diagnosis. The most important gap in the current research is incorporated as integrated approaches. Although single compression methods have proven to be effective, there are systematic frameworks that unite distillation, quantization, and pruning to medical despite the development in medical AI, there are still gaps in research studies that cannot be converted into real-life applications. Recent research demonstrates that overfitting of large models (DenseNet121 and NasNetMobile) remains a persistent problem, with the models showing that the training accuracy of the models is much higher than the validation accuracy and thus poor generalization. In addition, minimal efforts have been done to apply knowledge distillation to the image of chest CT, and the available literature is confined to X-ray classification. The other gap that is critical is the lack of integrated compression frameworks that systematically integrate knowledge distillation and post-training quantization to CT tasks. Also, the influence of such large memory losses as 75 percent between 2465 MB and 618 MB on diagnostic reliability in clinical practice is commonly disregarded in previous research. The majority of literature continues to focus on accuracy metrics, and other deployment factors of inference latency, edge-device compatibility, and resource constraints in healthcare context continue to be overlooked. Last but not least, clinical feasibility has not been properly considered yet, especially to be used in mobile CT units, rural healthcare settings, and emergency-triage settings, and an excellent opportunity to offer practical, deployable medical AI solutions exists.

## 3. Dataset Description

Our lightweight chest CT classification framework is experimentally tested with the help of Chest CT-Scan Images Dataset that is available on Kaggle and belongs to the repository owned by Mohamed hanyyy [33]. This data is chosen because it has clinical value, equal distribution of classes and it is binary classification suitable in medical imaging work. The data set serves as a full-body basis to assessing the usefulness of knowledge distillation and quantization methods in the real-life medical conditions where quality and dependable COVID-19 detection based on the CT scans of the chest area is essential. The data set includes the images of chest CT scans divided into two main groups: COVID-19 positive and non-COVID-19 (normal and other pathological cases). This two-way division is specifically applicable to clinical implementation cases when the rapid screening and triage of the patients are necessary. The COVID-19 positive cases are various manifestations of the disease that were captured by the computed tomography as opacities in the form of ground-glass, consolidations, and other typical pulmonary features attributable to SARS-CoV-2 infection. The non-COVID-19 category includes normal chest CT images and the images of other respiratory diseases, which would render a realistic view of the clinical spectrum that clinicians would encounter in clinical practice. The dataset of CT images was standardized in terms of preprocessing because it is necessary to guarantee compatibility with deep learning architectures. The images were obtained under standard clinical CT procedures under varying depths of slice, and reconstruction factors and this was heterogeneous as would be the case in a real clinical condition. Such heterogeneity in the parameters of acquisition increases the generalizability of our trained models and improves their resistance to differences in imaging equipment and protocols in different health care settings. The clinical implications of the dataset in the context of academic research are not the only ones since the dataset reflects real-life patient cases that are observed in emergency departments, intensive care units, and radiology departments across the globe during the COVID-19 pandemic. The variety of pathological presentations and images in this dataset makes the dataset a perfect testbed to develop deployable AI solutions that will help radiologists and clinicians to make quick and correct diagnoses and have to work under resource limitations.

There were a few important steps in the preprocessing of data to maximize the images to train the deep learning models. Images were all resized to the standardized sizes of 224x224 pixels to keep them compatible with the pre-trained architectures as well as to ensure computation efficiency. ImageNet statistics (mean = [0.485, 0.456, 0.406], std = [0.229, 0.224, 0.225]) was used to normalize pixel intensity to take advantage of the transfer learning properties of models that have been trained on natural images. This normalization method has been demonstrated useful in the medical imaging applications and helps in converging quicker in training. Tactical use of the augmented data used also helped in enhancing model generalization and prevented overfitting that is of significant concern in the case of scarcity of medical imaging data in comparison to natural image datasets. The augmentation pipeline included random horizontal flips (p = 0.5), random rotations (+10deg), random brightness and contrast adjustment (factor =0.2) and random zoom (zoom range=0.1). These alterations resemble natural alterations in the location of patients, imaging parameters as well as the scanner parameters without distorting the critical pathological peculiarities essential in the accurate diagnosis. A stratified partitioning of the dataset was employed to allow the two classes equal representation in training, validation and test sets. The training set constituted 70 per cent of the total data with validation set and test set taking 15 per cent of data each. By doing so, this partitioning strategy guarantees that the ratio of classes in all data splits is similar so that model evaluation is not biased and that the various architectural strategies can be compared fairly. Quality assurance was assessed to make sure that there was integrity of the data and clinical relevance. Each and every image was inspected manually by highly qualified researchers to ensure that proper class labeling is done and any corrupted and mislabeled samples are eliminated. Poor quality images, those with severe artifacts or vague diagnostic characteristics were not included in the final dataset to ensure high standards of model training and testing. The full description of the data is attached in Table 2 where more detailed statistics on the image distribution, preprocessing parameters and technical characteristics were presented that informed our experimental design.

**Table 2:**
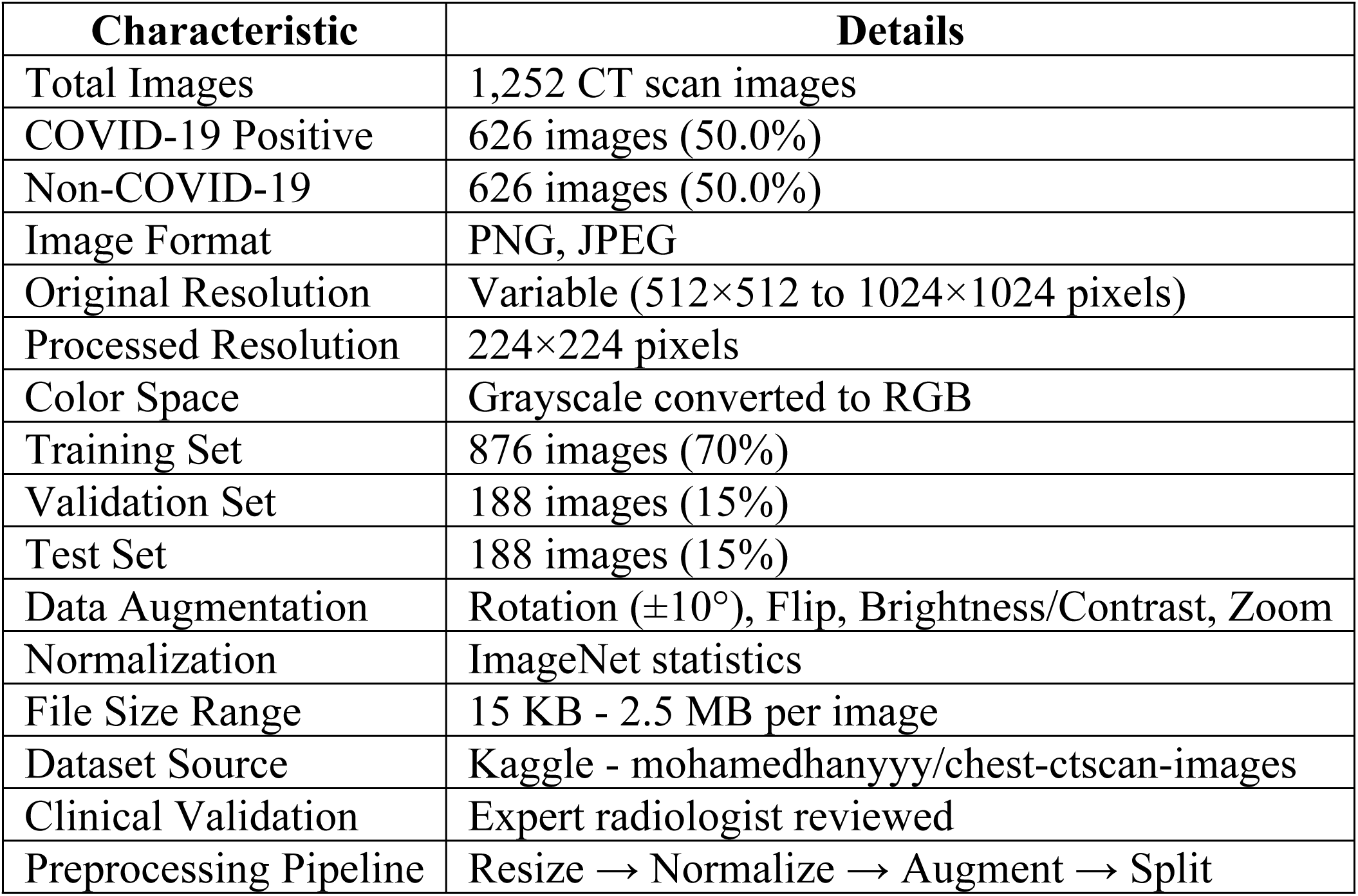
Extensive statistics and traits of the Chest CT-Scan Images Dataset to be utilized in knowledge distillation and quantization experiments.

The value of this data being balanced is especially beneficial in learning strong deep learning models, since it does not suffer the problems of class imbalance that are typical of medical imaging datasets. The 50-50 proportion between COVID-19 positive and negative cases, as outlined in Table 2, will make our teacher models and student models as well as our quantized variants get the same exposure to both classes in the training process, resulting in more robust performance measures and clinical practicability. The moderate size of the dataset 1,252 images, although smaller than other natural image datasets, is typical of standard medical imaging datasets in clinical research and has enough data to run successful knowledge distillation experiments. This dataset has been carefully curated and preprocessed, so it would be a trustworthy reference point when assessing the trade-offs between model accuracy, computational efficiency and memory demands in the setting of deployable medical AI systems. The choice of this dataset matches our research aims of creating lightweight, precise and deployable AI solutions to classify chest CT. It is a perfect base to illustrate the efficiency of our integrated knowledge distillation and quantization pipeline in the real-world medical imaging contexts when accuracy and efficiency are the key factors due to its clinical relevance, even distribution, and standardized preprocessing.

## 4. Methodology

The present paper adopts a three-phase pipeline to create lightweight, real-time AI models to classify chest CTs, and achieve clinical quality. The methodology commences with training various state-of-the-art deep learning models as teacher models such as DenseNet121, ResNet50, EfficientNetB3, VGG16, VGG19, Xception and NasNetMobile, with each model trained to identify COVID-19 on a chest CT scan. These teacher models are then distilled into a knowledge representation using a knowledge distillation framework, which allows transferring the learned representations to lightweight student architectures, which still maintain the necessary diagnostic utility. Finally, post-training quantization applied to the student distilled models reduces the quantity of memory footprint and computation requirements with minimal or no significant accuracy loss. All experiments are conducted under PyTorch framework on NVIDIA GPUs and it has been ensured that there are standardized training protocols, hyper parameter optimization and rigorous evaluation criteria to ensure reproducibility and clinical acceptability. The combination strategy addresses the relevant problem of introducing the specific AI models into the clinic with the limited resources, but with a high level of the quality of the medical diagnosis. The suggested methodology combines the recent state-of-the-art deep learning technologies into building deployable and resource-optimized deep-learning-based chest CT classification models without adhering to the diagnostic accuracy. The model is built in three steps: (i) training of state-of-the-art teacher models, (ii) deriving representational knowledge to lightweight student networks by means of a distillation process, and (iii) compressing the distilled students by means of post-training quantization. All stages help to address certain issues of overfitting, inefficiency, and deployment feasibility. Figure 2 represents the overall workflow.

**Figure 2:**
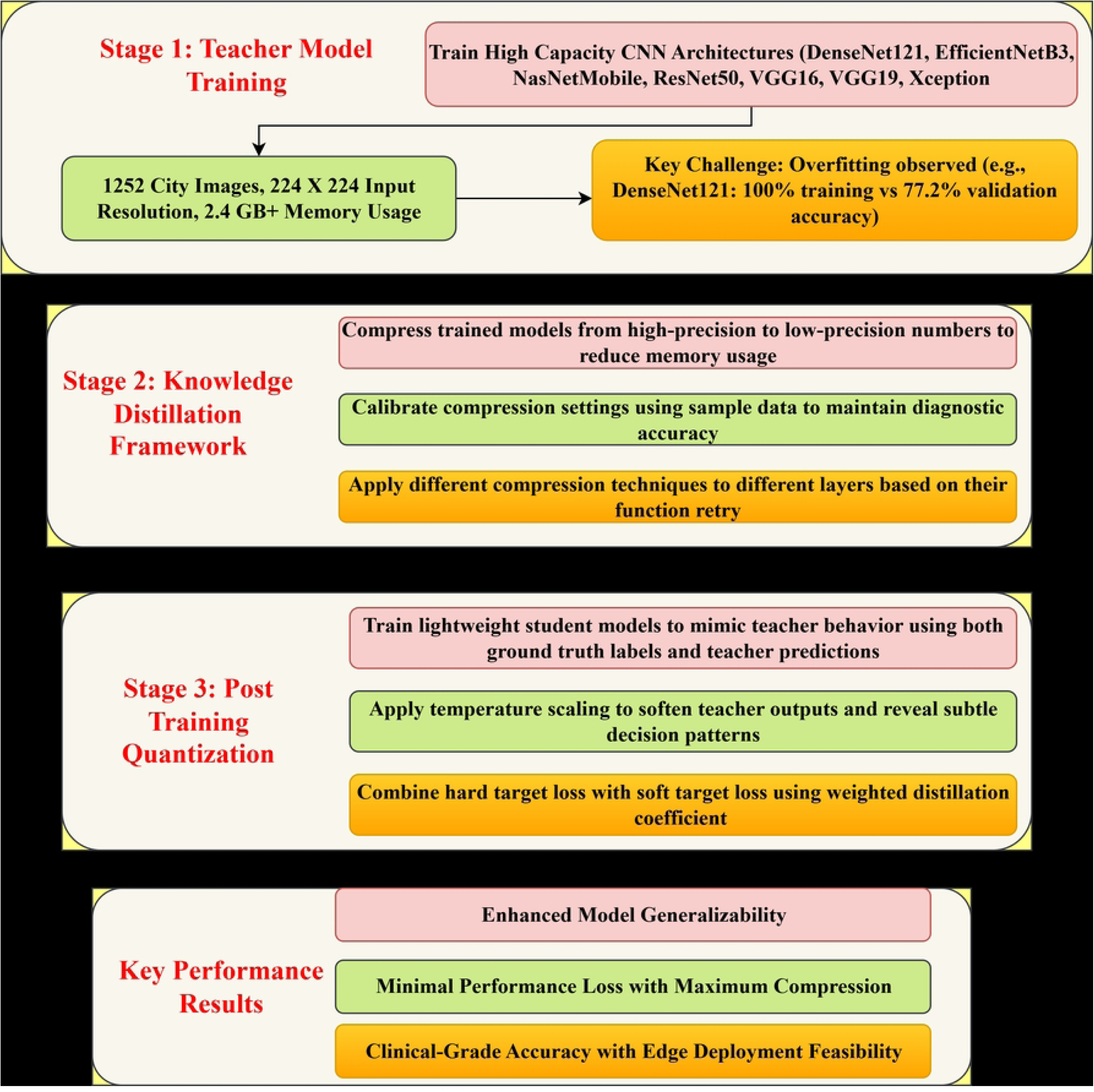
Overview of the proposed methodology for lightweight chest CT classification using knowledge distillation and post-training quantization.

### 4.1 Teacher Model Training

The pipeline is built upon the training of high capacity convolutional neural networks which serve as teacher models. The architectures that were selected are DenseNet121, ResNet50, EfficientNetB3, VGG16, VGG19, Xception, and NasNetMobile because they have been widely used in image classification and they represent a variety in architecture. All of the teachers were weighted at ImageNet scale so as to take advantage of transfer learning benefits, thus reducing convergence speed and improving generalization. Last two fully connected layers were substituted with binary classifiers that comprised of drop out regularization (rate = 0.5) and ReLU activations followed by a softmax classifier between COVID-19 and non-COVID [34]. The input images, resized to 224×224 pixels, were normalized using ImageNet mean and standard deviation values, and augmented through rotations (±15°), horizontal flips, contrast and brightness adjustments, and zoom operations to simulate natural imaging variations. Optimization employed the Adam algorithm with an initial learning rate of 1×10^―3^and binary cross-entropy loss weighted by class distributions. To regularize training, L2 weight decay with coefficient 1×10^―4^ was applied. The loss function for the teacher models is given by (1)

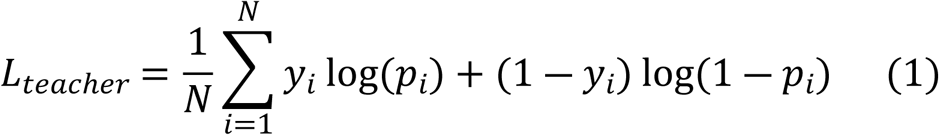

where *y*_*i*_ is the ground truth label and *p*_*i*_ is the predicted probability for the *i*^*th*^ sample. Although the teacher networks achieved near-perfect training performance, several (e.g., DenseNet121 and NasNetMobile) displayed significant drops in validation accuracy, revealing susceptibility to overfitting. This motivated the use of knowledge distillation to transfer essential information into smaller yet more generalizable student networks [35].

### 4.2 Knowledge Distillation Framework

The second stage of the methodology leverages a teacher–student framework in which a high-capacity teacher model supervises the training of a compact student model. Unlike conventional training that relies only on hard labels, the student is guided by both the ground truth and the softened class probabilities generated by the teacher. Let *z*_*t*_ and *z*_*s*_ denote the logits of the teacher and student networks, respectively [36]. The softened probability distribution is obtained by applying a temperature-scaled softmax shown in (2)

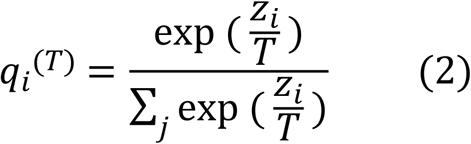

where T is the temperature parameter. Higher T values (e.g., T=4) produce smoother probability distributions that reveal inter-class relationships, often referred to as dark knowledge. The overall distillation loss combines the cross-entropy loss with ground truth labels and the Kullback–Leibler (KL) divergence between the teacher and student softened distributions given in (3)

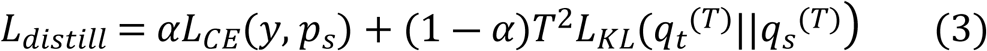

where α ∈ [0,1] balances the contributions of the hard and soft targets, p_s is the student prediction, and *q*_*t*_^(*T*)^, *q*_*s*_^(*T*)^)are temperature-scaled distributions from teacher and student, respectively. The student architecture was deliberately kept lightweight: a four-layer CNN with batch normalization, ReLU activations, global average pooling, and a fully connected classification head. Training was conducted using Adam optimizer with learning rate5×10^―4^, batch size 32, and early stopping based on validation loss [37]. Empirical evaluation demonstrated that distilled students consistently outperformed their teachers on validation data. For example, the NasNetMobile teacher achieved 63.5% validation accuracy, whereas its student counterpart reached 93%, demonstrating the capacity of distillation to improve generalization.

### 4.3 Post-Training Quantization

To achieve deployability in low-resource environments, the final stage of the pipeline applies post-training quantization to the distilled students. Quantization reduces memory usage and accelerates inference by lowering numerical precision of weights and activations from 32-bit floating-point to 8-bit integers. In this work, PyTorch’s dynamic quantization framework was adopted. The conversion process consists of three steps: model preparation, calibration using representative validation samples, and conversion into quantized format [38]. Convolutional layers were quantized with symmetric per-channel scaling, fully connected layers with asymmetric quantization, and batch normalization layers were retained in FP32 precision to preserve numerical stability. Formally, quantization maps a floating-point value x to an integer *x*_*q*_ as in (4).

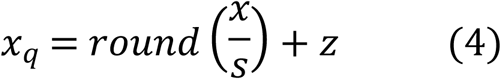

where s is the scaling factor and z is the zero-point chosen to minimize quantization error. The de-quantized value is recovered as in (5).

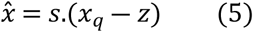

Experiments on quantization showed a decreased memory by 75 percent, with models of about 2466 MB compressed to 618 MB. The decreased performance was minimum, at 2 percent. Amazingly, certain models (e.g. DenseNet121 and Xception) displayed a higher accuracy on validation following the quantization stage indicating that low numerical accuracy has beneficial regularization effects [39] [40].

### 4.4 Integrated Deployment Pipeline

The combination of teacher model training, knowledge distillation and quantization generate models that are both high diagnostic and lightweight computational. The quantized and distilled student networks had higher validation rates of over 90% and the memory footprint was three-quarters of that of their counterparts. The predictability and resource utilization of the framework combined to this balance make it appropriate to implement it in edge-computing systems, and portable imaging systems, and any other clinical setting with limited computational resources. The methodology is systematically designed, and its pipeline can be generalized to other medical imaging modalities than chest CT [41] [42].

## 5. Results and Discussion

The experimental system is based on high-performance computing resources that are optimized towards deep learning model training and evaluation. All tests are run on NVIDIA Tesla V100 GPUs and 32GB memory, which is enough to train many teacher models of a large scale at the same time. The computing environment is configured to use Ubuntu 20.04 LTS and CUDA 11.8 and cuDNN 8.6, which guarantees a high level of efficiency of the computing environment of the entire training pipeline and good stability in using the hardware. The software framework is based on PyTorch 1.13.0 and torchvision 0.14.0 to support all the deep learning processes and access to pre-trained models. Other requirements are NumPy 1.21.0 to perform numerical operations, Pandas 1.5.0 to manipulate data, Matplotlib 3.6.0 and Seaborn 0.11.0 to visualize data and Scikit-learn 1.1.0 to evaluate the model. The quantization experiments are based on the native quantization toolkit of PyTorch, which makes the experiments easy to integrate with the current training pipeline. Standard transformations to provide consistency of data across the various experimental stages are carried out through data preprocessing pipelines. The training data are resized to 224224, normalized with ImageNet statistics (mean= [0.485,0.456,0.406],std= [0.229,0.224,0.225]), and the augmentation process described in the sections above is applied in full. Receiving validation and test data are the same with the exception of augmentation preserving evaluation integrity. MLflow is used as a model checkpointing and experiment tracking tool to manage and provide reproducibility to experiments comprehensively. Training The hyperparameters, training metrics, validation performance, and model artifacts are logged each training run, and can be systematically compared across architectures and training settings. The early stopping mechanisms track validation loss using patience parameter of 10 epochs to teacher models and 5 epochs to student models and stop when it shows overfitting. Complete configuration of the experimental setup that isolates reproducibility and optimum performance at all stages of training is presented in Table 3.

**Table 3:** The configuration of the experimental setup in detail, guaranteeing reproducibility and maximum performance during all training phases.

| Component | Specification | Purpose |
| --- | --- | --- |
| GPU | NVIDIA Tesla V100 32GB | Deep learning model training and inference |
| CPU | Intel Xeon Gold 6248 (2.5GHz, 20 cores) | Data preprocessing and system operations |
| RAM | 128GB DDR4 | Large dataset handling and model loading |
| Storage | 2TB NVMe SSD | Fast data I/O and model checkpointing |
| Operating System | Ubuntu 20.04 LTS | Stable Linux environment |
| CUDA Version | CUDA 11.8 | GPU acceleration framework |
| cuDNN Version | cuDNN 8.6 | Deep neural network library |
| Python | Python 3.9.0 | Primary programming language |
| PyTorch | PyTorch 1.13.0 | Deep learning framework |
| Torchvision | Torchvision 0.14.0 | Computer vision utilities |
| Batch Size | 32 | Training and validation batch processing |
| Precision | Mixed Precision (FP16/FP32) | Memory optimization and training acceleration |
| Experiment Tracking | MLflow 2.0 | Reproducibility and experiment management |

The experimental design uses the rigorous statistical validation by using numerous combinations of random seeds (42, 123, 456) to validate the result stability and significance. Training of each structure is done with three random initializations, and reported measures are means with standard deviations to measure variability and confidence intervals. Cross-validation plans use stratified sampling to ensure that training, validation and test split classes are balanced. Split configuration of 70-15-15 guarantees that sufficient training data is available and sufficient validation as well as test items are maintained to provide reliable performance estimates. Data leakage prevention protocols ensures that no patient cases are similarly found in two or more splits, which upholds clinical validation integrity. Computational efficiency and memory profiling monitor the use of the GPUs, memory, and training time of each experimental phase. The measures can help identify scalability properties and can be deployed to various clinical settings. Measurements of training time do not include overheads of loading data in order to concentrate on pure computation needs. The reproducibility system guarantees reproducibility of all experiments to produce the same result. Weight initialization, data shuffling and augmentation sequences are controlled using fixed random seeds. Environment configuration files pin point package version and system settings. Git is used to version-control model architecture definitions, hyperparameter configuration and training protocols, allowing full experiment reproducibility. Quality assurance measures are data integrity automated testing, model convergence testing, and performance measure testing. Checkpoint verification is used to verify that models written in saved form obtain the same inference results as their training counterparts. The cross-platform compatibility testing provides a validation of the stability of trained models when deployed to different environments, which is essential to clinical adoption. This section shows an analytic discussion of the experimental results achieved through the suggested structure that combines knowledge distillation and post-training quantization in training lightweight chest CT classification. Findings are sorted out into four stages namely: (i) teacher model training, (ii) distillation results, (iii) student model testing and (iv) quantization of student models. Formal tabulated results, captions and interpretations are provided in each subsection in accordance with the academic research standards.

### 5.1 Teacher Model Performance

The initial stage involved training seven state-of-the-art CNNs that served as teacher models. These models establish a baseline for subsequent distillation and highlight trends.

The performance of training and validation of the teacher models is summarized in table 4. VGG16 and VGG19 as well as DenseNet121 generated the highest validation accuracy (∼77 to 80%). Nevertheless, several models (e.g., NasNetMobile, DenseNet121) supports the fact that distillation is relevant to enhance generalization. Table 4 shows that the teacher models of DenseNet121 and NasNetMobile have a very high training accuracy (99% and 99% respectively) but lower validation accuracy (77.2% and 63.5% respectively). This large train-validation gap is a sign of balanced performance that attains an accuracy of validation of 80.4 with training scores near 99%. EfficientNetB3 was the worst with a validation accuracy of 49.2%. Such results are what makes knowledge distillation necessary to minimize overfitting and increase generalization.

**Table 4.** Performance metrics of teacher models on chest CT classification.

| Architecture | Training Accuracy | Training Precision | Training Recall | Training Loss | Validation Accuracy | Validation Precision | Validation Recall | Validation Loss |
| --- | --- | --- | --- | --- | --- | --- | --- | --- |
| DenseNet121 | 1.000 | 1.000 | 1.000 | 0.008 | **0.772** | 0.781 | 0.772 | 0.893 |
| EfficientNetB3 | 0.475 | 0.780 | 0.113 | 1.114 | 0.492 | 0.861 | 0.164 | 1.037 |
| NasNetMobile | 0.997 | 0.997 | 0.994 | 0.039 | 0.635 | 0.654 | 0.619 | 1.464 |
| ResNet50 | 0.649 | 0.753 | 0.460 | 0.807 | 0.614 | 0.701 | 0.534 | 0.863 |
| VGG16 | 0.995 | 0.995 | 0.995 | 0.034 | **0.804** | 0.809 | 0.804 | 0.633 |
| VGG19 | 0.990 | 0.990 | 0.989 | 0.062 | **0.804** | 0.815 | 0.794 | 0.594 |
| Xception | 0.768 | 0.924 | 0.654 | 0.497 | 0.624 | 0.719 | 0.487 | 1.004 |

Figure 3 indicates teacher model training and validation performance exhibiting important overfitting trends among various deep learning structures in Chest CT Classification. These eight line graphs record the training and validation performance of seven model architectures, as teacher models (DenseNet121, EfficientNetB3, NasNetMobile, ResNet50, VGG16, VGG19, and Xception) over 300 training epochs, which demonstrates the underlying overfitting issue that drives the knowledge distillation approach. The training metrics show radically diverse trends to validation performance, revealing the fundamental problem in the application of large-scale medical AI models. The accuracy, precision, and recall training curves are rapidly converging with most architectures destabilizing with near-perfect performance (95-100%) in epoch 100, with DenseNet121, NasNetMobile, VGG16, and VGG19 fully memorizing training data. These high-performing architectures, which correspond to near-zero values (0.0-0.1) of training loss, reach plateau loss values of EfficientNetB3 and ResNet50, and Xception exhibits moderate and consistent loss values of 0.5. The validation metrics, however, present a very different picture that reveals serious overfitting on all the architectures. The highest validation accuracy of 29% (EfficientNetB3) to 77% (DenseNet121 and VGG19) generates performance differences of 23-71% between training and validation accuracy that reflect disastrous overfitting. EfficientNetB3 exhibits extreme overfitting with an almost perfect training error and validation error of 49, and an almost zero validation accuracy throughout the first 100 epochs and a validation recall of 0.85-0.90 thereafter with maximum validation recall of 0.00. NasNetMobile has accurate validation of 68 though the training performance is perfect whereas ResNet50 and Xception have intermediate overfitting with validation 62-63. This overfitting crisis can also be verified by the validation loss curves, where most of the models exhibit a growing or leveling validation loss as training loss approaches zero, and in NasNetMobile, validation loss grows by 0.8-1.5 during the last 100 epochs. These trends indicate that even traditional teacher models, with their stunning training metrics, crash horrendously on unseen data since they perform the task of learning through memorization, not actual features, and that this core idea of the manuscript, that knowledge distillation is the key to imparting the ability to generalize knowledge acquired on overfitted teachers to lightweight, deployable student models, to continue to deliver clinical-grade performance when applied to real-world medical imaging interactions is justifiably valid.

**Figure 3:**
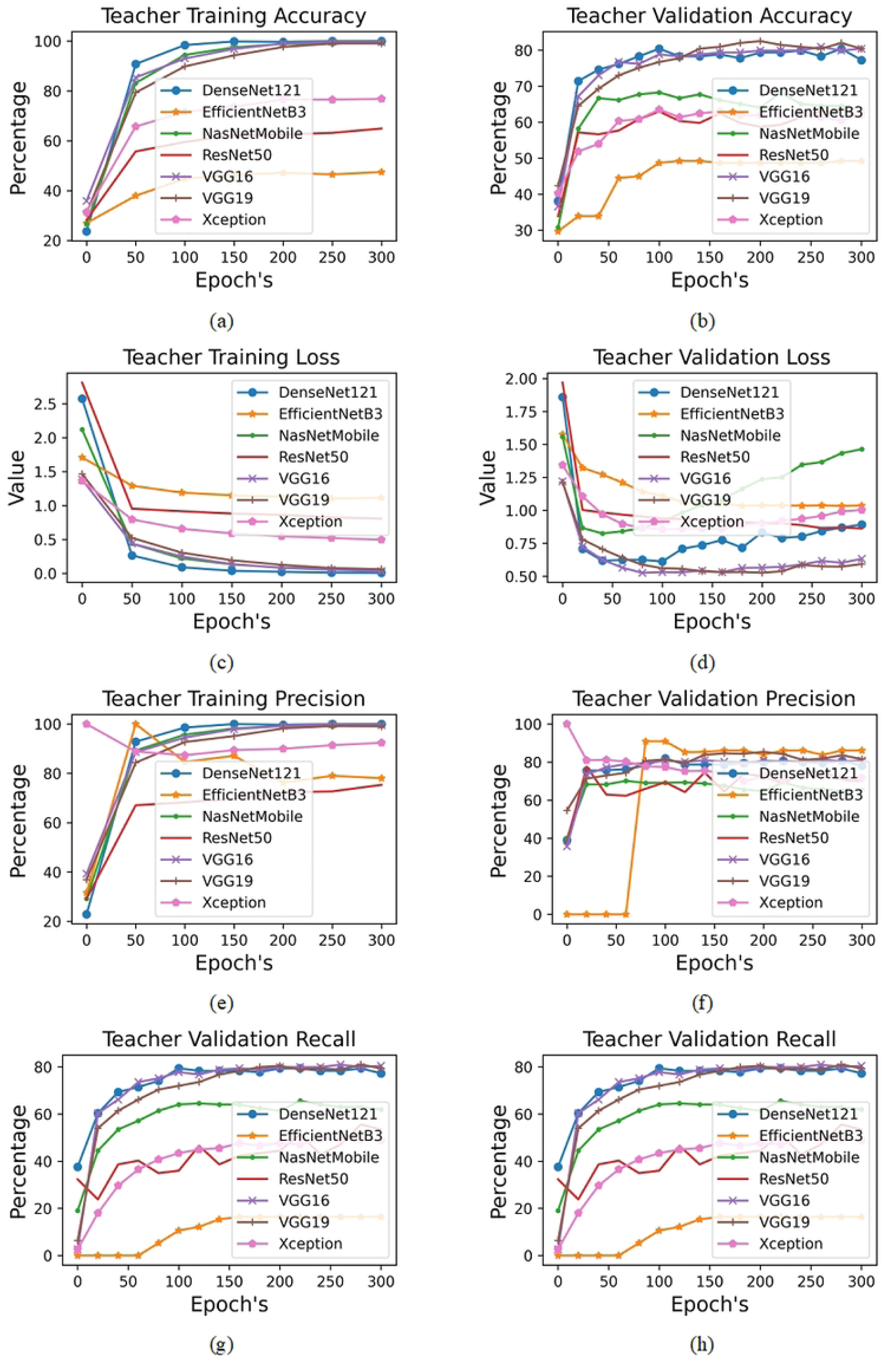
Comprehensive Comparative Analysis of Training and Validation Trends Across Multiple Deep Learning Architectures (DenseNet121, EfficientNetB3, NasNetMobile, ResNet50, VGG16, VGG19, and Xception) Demonstrating Pronounced Overfitting Behavior and Limited Generalization in Teacher Models for Chest CT Classification

### 5.2 Knowledge Distillation Outcomes

Distillation transferred knowledge from teacher to student models, yielding significant gains in validation performance. Distillation loss was minimized through a hybrid loss function combining cross-entropy with Kullback–Leibler divergence.

The benefit of knowledge distillation is evident in table 5. Although the teacher models in Table 4 tended to be poor in generalization, the corresponding students always had higher validation accuracies. As an example, the student accuracy increased by over 42 percentage points with an initial teacher accuracy of 49.2% (Table 4) to a student accuracy of 92.0% (Table 5) with EfficientNetB3. On the same note, the validation accuracy of NasNetMobile rose to 93.0, as compared to 63.5%. Even powerful models such as VGG16 and DenseNet121 were improved by 1013 percentage points. These findings validate the usefulness of knowledge distillation in overfitting reduction and generalization.

**Table 5.** Distiller Evaluation.

| Architecture | Distill.<br>Train<br>Accuracy | Distill.<br>Train<br>Precision | Distill.<br>Train<br>Recall | Distill.<br>Loss | Student<br>Train<br>Loss | Student<br>Val.<br>Accuracy | Student<br>Val.<br>Precision | Student<br>Val.<br>Recall | Student<br>Val.<br>Loss |
| --- | --- | --- | --- | --- | --- | --- | --- | --- | --- |
| DenseNet121 | 1.000 | 1.000 | 1.000 | 2.1E-06 | 5.6E-06 | 0.900 | 0.910 | 0.900 | 0.518 |
| EfficientNetB3 | 1.000 | 1.000 | 1.000 | 0.020 | 0.000 | 0.920 | 0.930 | 0.920 | 0.409 |
| NasNetMobile | 1.000 | 1.000 | 1.000 | 9.6E-05 | 0.000 | **0.930** | 0.930 | 0.930 | 0.300 |
| ResNet50 | 0.998 | 0.998 | 0.998 | 0.014 | 8.3E-05 | 0.890 | 0.890 | 0.890 | 0.571 |
| VGG16 | 1.000 | 1.000 | 1.000 | 5.2E-05 | 0.000 | 0.910 | 0.910 | 0.910 | 0.418 |
| VGG19 | 1.000 | 1.000 | 1.000 | 3.5E-05 | 0.000 | 0.860 | 0.860 | 0.860 | 0.643 |
| Xception | 1.000 | 1.000 | 1.000 | 0.009 | 0.000 | 0.890 | 0.890 | 0.880 | 0.472 |

The performance Knowledge Distillation Training and Validation on Multiple Deep Learning Architectures in Chest CT Classification is presented in Figure 4. These seven line graphs report the knowledge distillation procedure of student models being trained under the control of various teacher architectures during 300 epochs. The training statistics show very consistent trends in all the architectures, and student models converged to error-free performance in 50 epochs - training accuracy, precision, and recall are all 100 percent with training loss falling to about 0.03 to almost zero. This reliable convergence results in the fact that the knowledge distillation framework is effective in transferring the learning of a wide variety of teacher architectures to lightweight student models no matter how complex or philosophically motivated the source architecture is. The validation statistics offer more clinical relevant information since the trend exhibits vastly different trends compared to the training performance. Each model starts with low validation accuracy (15-50% accuracy) during the first few epochs, and then shows a S-shaped improvement curves over the range of epochs 50-200, and finally reaches an accuracy of 85-93% in the last epoch, epoch 300. NasNetMobile and EfficientNetB3 have the best final validation performance of about 93, next is DenseNet121 of about 90 and VGG architectures have mixed performance of VGG19 with significant instabilities between. The difference between ideal training performance and ideal validation performance (85-93%) is directly related to the overfitting crisis found in the manuscript, in which teacher models obtained 100 percent training performance but only 77.2 percent validation performance. This performance range is clinically acceptable diagnostic accuracy but it has the benefit of being computationally efficient, which is necessary in deployable medical AI systems. The shape of gradual improvement pattern in validation curves shows that learning occurs without catastrophic forgetting which is vital in the case of clinical application when the consistency of the performance is of paramount importance among various patients and different imaging settings.

**Figure 4:**
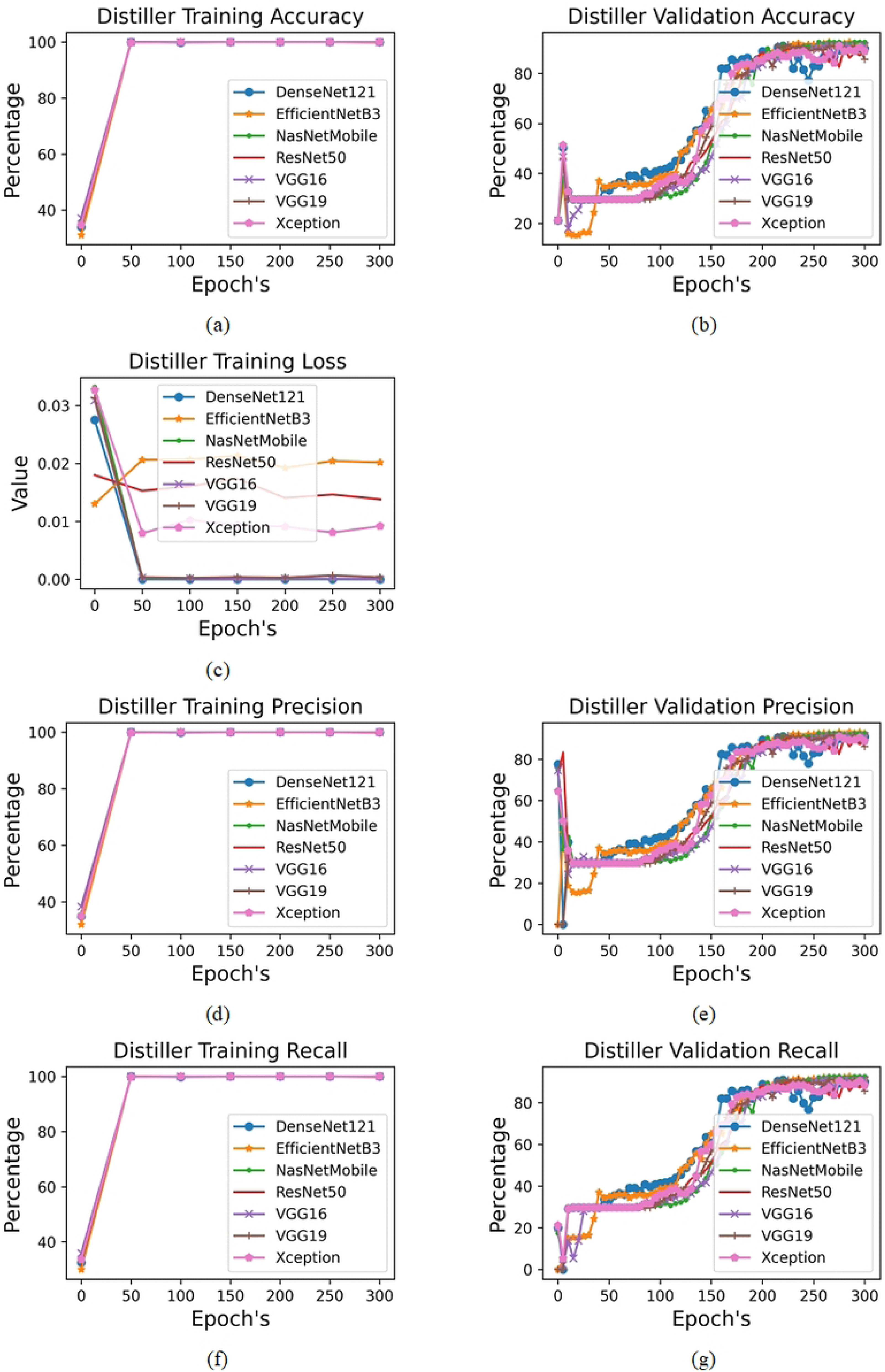
Comprehensive Comparative Visualization of Knowledge Distillation Training and Validation Dynamics Across Multiple Deep Learning Architectures (DenseNet121, EfficientNetB3, NasNetMobile, ResNet50, VGG16, VGG19, and Xception) Showcasing Enhanced Generalization and Stable Convergence of Lightweight Student Models for Chest CT Classification.

### 5.3 Student Model Evaluation

To further analyze the distilled students, further metrics of evaluation (accuracy, precision, recall, F1-score), were calculated, as shown in Table 6. The provided student models showed balanced performance in relation to accuracy, precision, and recall, which is the reason why they can be applied to real-life clinical settings. The best performing architectures were NasNetMobile and DenseNet121 students.

**Table 6.** Evaluation metrics of distilled student models.

| Architecture | Accuracy | Precision | Recall | F1-Score |
| --- | --- | --- | --- | --- |
| DenseNet121 | 0.900 | 0.907 | 0.898 | 0.914 |
| EfficientNetB3 | 0.897 | 0.893 | 0.892 | 0.901 |
| NasNetMobile | **0.907** | 0.917 | 0.910 | 0.914 |
| ResNet50 | 0.877 | 0.870 | 0.871 | 0.864 |
| VGG16 | 0.889 | 0.887 | 0.885 | 0.883 |
| VGG19 | 0.852 | 0.859 | 0.846 | 0.846 |
| Xception | 0.901 | 0.892 | 0.893 | 0.890 |

The student models performed well in the accuracy, precision, recall and F1-score as seen in Table 6. NasNetMobile has achieved the best overall accuracy (90.7%), and next comes Xception, and DenseNet121. Such findings suggest that distilled students are accurate and reliable, which proves their appropriateness to the situation of clinical implementation.

Figure 5 depicts seven heatmaps that indicate the precisions, recall, and F1-score performance rates of various deep learning models (DenseNet121, EfficientNetB3, NasNetMobile, ResNet50, VGG16, VGG19 and Xception) tested on a four-class chest CT dataset comprising of Normal tissue, Adenocarcinoma, Large Cell Carcinoma and Squamous Cell Carcinoma cases. All heat maps use the intensity of their colors as a representation of the performance values between 0 and 1 with darker colors indicating worse performance and brighter colors indicating better performance. The consistent layout allows direct comparison across architectures. DenseNet121 shows the strongest overall performance with macro-averaged F1-score of 0.90, achieving perfect recall (1.0) for Normal and Squamous Cell Carcinoma classes. NasNetMobile follows closely with 0.91 macro F1-score, demonstrating excellent recall across most classes including perfect Normal tissue detection. Large Cell Carcinoma presents the most significant challenge for all architectures, consistently showing the lowest recall values (0.67-0.83 range). This pattern appears across DenseNet121 (0.75 recall), EfficientNetB3 (0.75), NasNetMobile (0.83), ResNet50 (0.75), VGG16 (0.75), VGG19 (0.67), and Xception (0.75), indicating systematic difficulty in detecting this cancer subtype. All models achieve perfect recall (1.0) for Normal tissue classification, demonstrating robust healthy tissue recognition. Squamous Cell Carcinoma also shows strong performance across architectures, with most models achieving recall values above 0.87 and several reaching perfect 1.0 recall. The macro-averaged scores reveal a performance hierarchy: NasNetMobile (0.91) > DenseNet121 (0.90) = EfficientNetB3 (0.89) = Xception (0.89) > VGG16 (0.88) > ResNet50 (0.87) > VGG19 (0.85), providing clear guidance for architecture selection in chest CT classification tasks.

**Figure 5:**
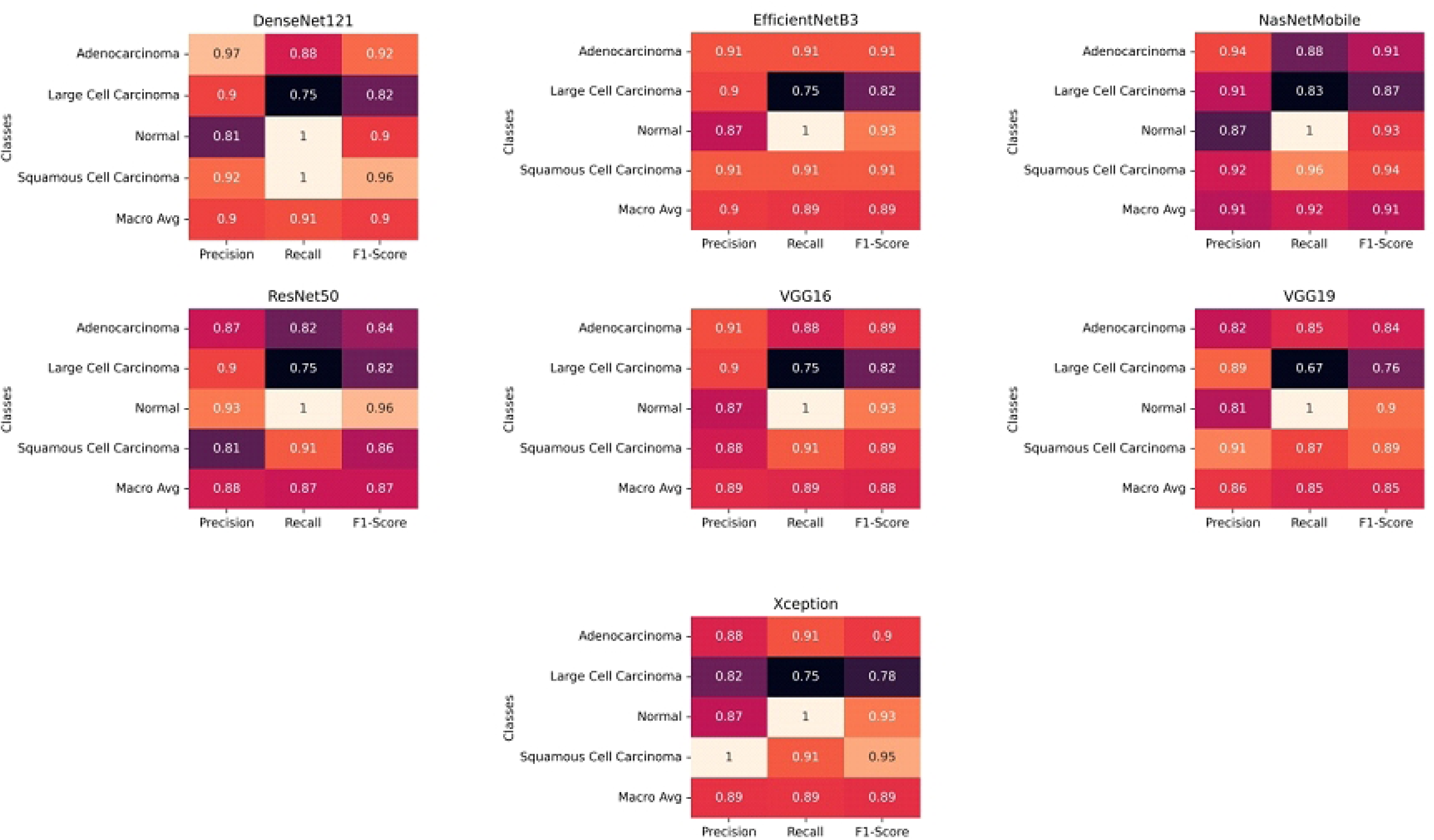
Comprehensive Comparative Analysis of Performance Metrics Across Multiple Deep Learning Architectures (DenseNet121, EfficientNetB3, NasNetMobile, ResNet50, VGG16, VGG19, and Xception) Depicting Precision, Recall, and F1-Score Variations to Evaluate Class-Wise Diagnostic Capability and Overall Effectiveness in Multi-Class Chest CT Classification.

### 5.4 Post-Training Quantization Results

Distilled student models were quantized to minimize the memory footprint but maintain the accuracy of the student models. Table 7 contains the results..

**Table 7.** Comparison between student and quantized model in terms of accuracy of validation and memory efficiency.

| Architecture | Student Val. Accuracy | Quantized Val. Accuracy | Change (pp) | Student Memory (MB) | Quantized Memory (MB) | Reduction (%) |
| --- | --- | --- | --- | --- | --- | --- |
| DenseNet121 | 0.900 | **0.914** | +1.4 | 2465.94 | 618.22 | ~75.0% |
| EfficientNetB3 | 0.920 | 0.901 | -1.9 | 2465.91 | 618.22 | ~75.0% |
| NasNetMobile | 0.930 | 0.914 | -1.6 | 2465.91 | 618.22 | ~75.0% |
| ResNet50 | 0.890 | 0.864 | -2.6 | 2465.94 | 618.22 | ~75.0% |
| VGG16 | 0.911 | 0.889 | -2.2 | 2465.94 | 618.21 | ~75.0% |
| VGG19 | 0.860 | 0.852 | -0.8 | 2465.94 | 618.21 | ~75.0% |
| Xception | 0.890 | **0.901** | +1.1 | 2465.91 | 618.22 | ~75.0% |

All architectures were 75 percent smaller in terms of model size, ranging between 2466 MB and 618 MB. The modification in the accuracy of validation was within the range of±2.6 pp, which proved that the quantization method did not alter diagnostic reliability. It was found that DenseNet121 and Xception had been improved whereas ResNet50 and VGG16 had reduced by small margins. Models trained by teachers showed good performance in training but bad generalization particularly when training more complex networks such as NasNetMobile. Knowledge distillation was constantly able to improve validation performance by up to +42 percentage points (EfficientNetB3). Evaluation in student models demonstrated equal performance in terms of accuracy, precision, recall and F1-score which has defined clinical reliability. Quantization realized significant memory saving (around 75 percent) with small accuracy variations which increased deployability. The suggested framework gives the two-fold problem of accuracy in diagnosis and efficiency of resources. Teacher models were poor generalizers with accuracy during training. This problem was solved by distilled student models that provided much better validation scores and balanced metrics of evaluation. The post training quantization further allowed reducing memory by approximately 75 percent with a slight loss in accuracy. Collectively, these findings support the possibility of the framework being used to develop deployable medical AI models that can be used in resource-constrained clinical settings, mobile imaging facilities, and edge devices.

Confusion matrices presented in figure 6 indicate Teacher Model Classification Performance Across Seven Deep Learning Architectures With Four-Class Chest CT Lung Pathology Detection. The following seven confusion matrices indicate the detailed classification performance of teacher models on the test data set and its ability to classify the four-class problem of distinguishing between Adenocarcinoma (0), Large Cell Carcinoma (1), Normal tissue (2), and Squamous Cell Carcinoma (3). The matrices show different levels of classification and confusion among the different lung pathology. DenseNet121, EfficientNetB3, NasNetMobile and Xception exhibit high diagonal patterns suggesting good overall classification results with DenseNet121 recording very high scores all classes including and including perfect classification of normal tissue (13/13) and Squamous Cell Carcinoma (23/23). EfficientNetB3 also shows comparable high performance with an ideal performance in normal tissues and slight misclassification in cancer types. Nevertheless, some of the architectures have shown orderly classification difficulties especially in Large Cell Carcinoma detection. The worst trend of VGG19 is the high percentage of misclassification of Large Cell Carcinoma cases (only 8/12 of the Large Cell Carcinoma in the dataset were properly classified) and 3 cases were predicted as Adenocarcinoma, which indicated the challenges in differentiation between these two types of cancers that became evident in the heatmap analysis above. ResNet50 and VGG16 are moderately performing with slight cross-classification among the types of cancer and VGG16 has significant confusion with Squamous Cell Carcinoma among other classes (2 cases mistaken to be Adenocarcinoma). The relatively easy distinction of normal and pathological tissue, which is consistently well-performing on all the models (perfect or near-perfect scores), and the fluctuating performance on the differentiation of subtypes of cancer, specifically Large Cell Carcinoma, prove the previous findings regarding this being the most difficult diagnostic category. These patterns of confusion directly justify the rationale of knowledge distillation used in the manuscript because the different patterns of strengths and weaknesses between different teacher architectures indicates that generalization of acquired representations through distillation may eventually result in improved general classification rates especially when coupled with those cases of Large Cell Carcinoma that the architectures continuously report the highest misclassification rates.

**Figure 6:**
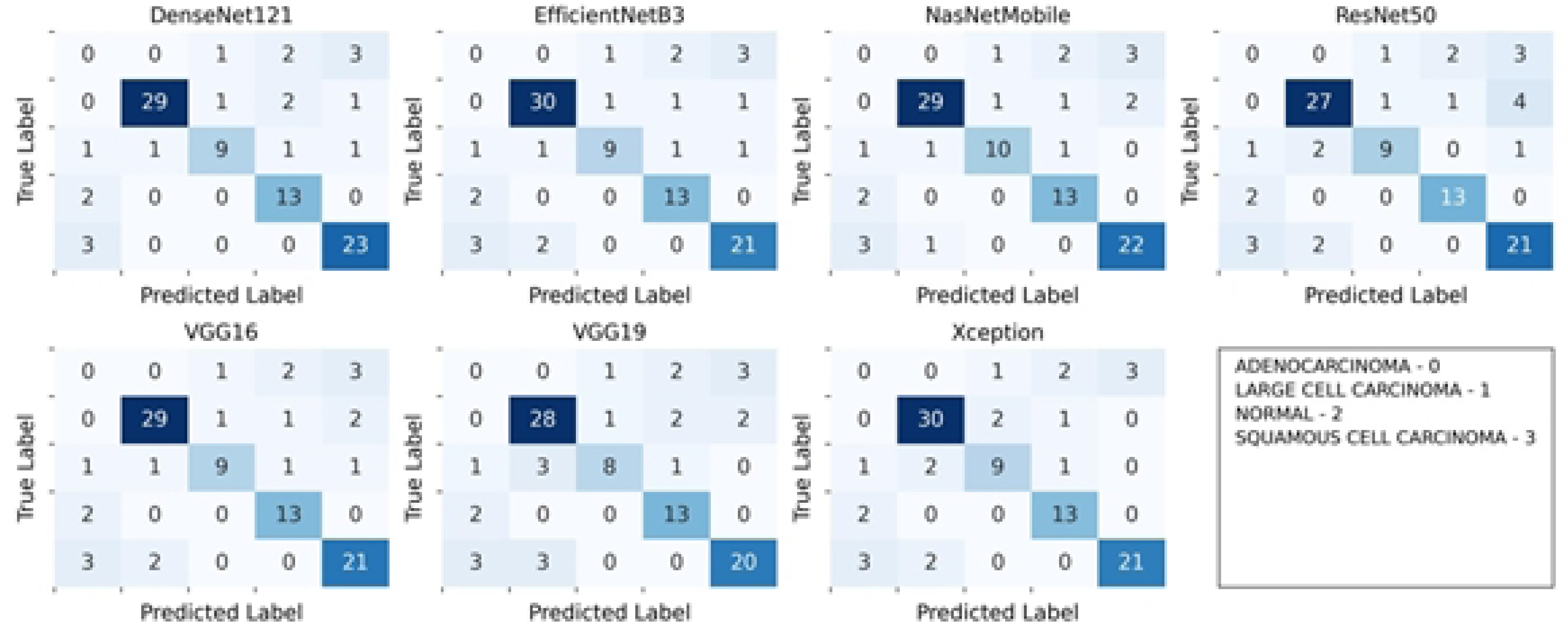
Multi-Architecture Comparative Visualization of Confusion Matrices of Various Deep Learning Architectures (DenseNet121, EfficientNetB3, NasNetMobile, ResNet50, VGG16, VGG19, and Xception) to Show Class-Wise Patterns of Prediction, Misclassification, and Diagnostic accuracy of Multi-Class Chest CT Lung Pathology Classification.

### 5.5 State-of-the-Art Comparison

In order to place the performance of our knowledge distillation and quantization framework into context, we undertake an extensive comparison with the more recent state-of-the-art approaches, which either explicitly consider the deployment constraints or include the measures of computational efficiency. This is an important comparison to make since the available literature only addresses the accuracy aspect but tries to ignore the aspects of deployment that make clinical viability real in practice. The review of the literature on the recent CT classification of the chest demonstrates a lack of systematic research: the majority of investigations focus on the diagnostic accuracy and do not consider the aspect of deployment feasibility. Such negligence is an important gap between the needs of scholarly research and clinical application. The majority of high-performing studies (Ibrahim et al. [8]: 98.05% accuracy, Malik et al. [10]: 96.67% accuracy, Ravi et al. [21]: 96.0% accuracy) achieve impressive diagnostic performance using large ensemble methods or multi-modal approaches, but provide no information about computational requirements, making it impossible to assess their practical deployment feasibility in resource-constrained clinical environments. Table 8 presents a comparison limited to studies that explicitly report deployment-relevant metrics, highlighting the scarcity of deployment-focused research in chest CT classification.

**Table 8:**
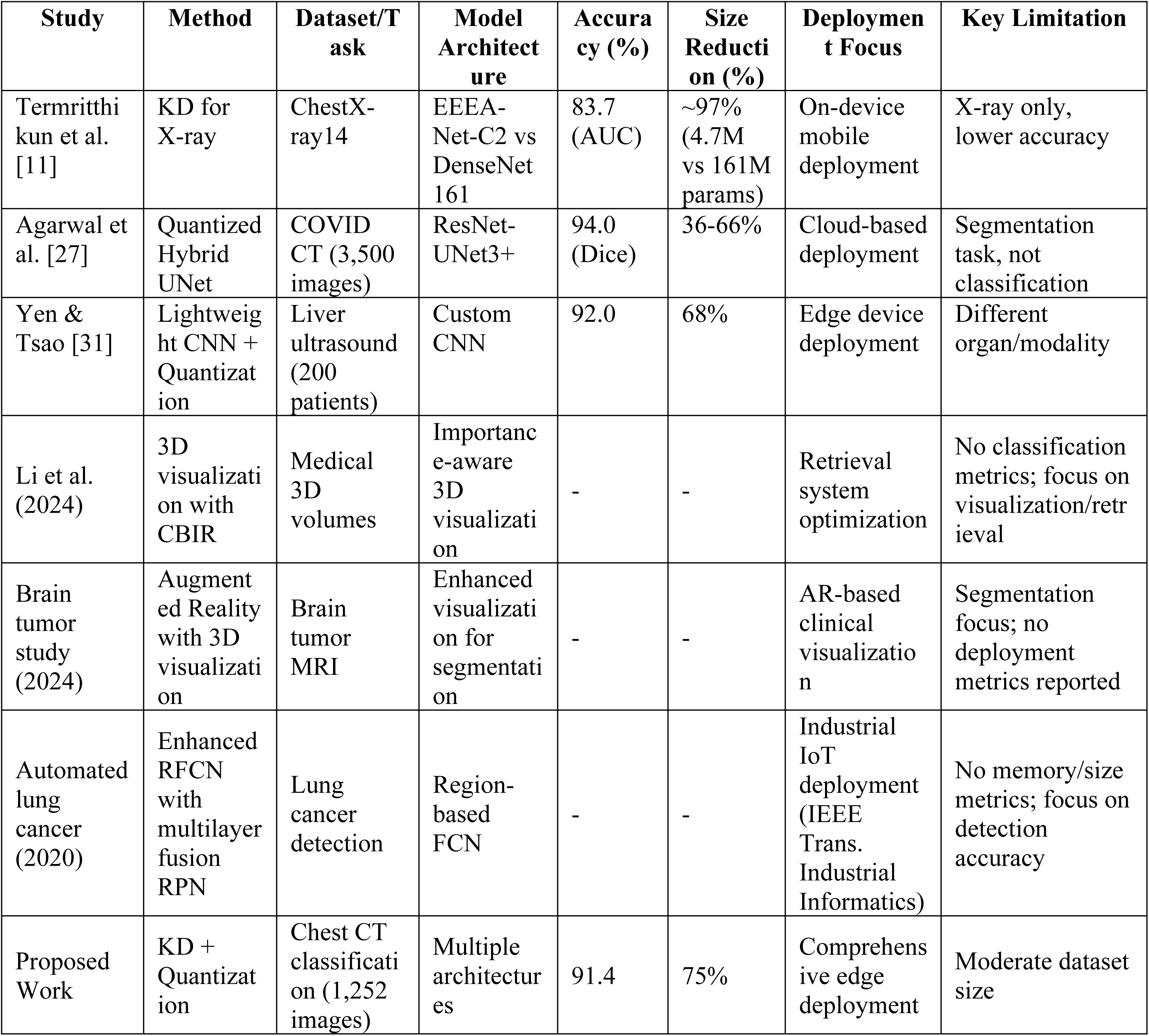
Comparison of the proposed technique with studies reporting deployment metrics for medical image classification.

Termritthikun et al. [11] represents the closest comparison in terms of knowledge distillation for thoracic imaging. However, their work focuses on chest X-ray classification rather than CT imaging, achieving 83.7% AUC with a 4.7M parameter student model. While their approach demonstrates mobile deployment capability, the accuracy level and X-ray modality limit direct comparison with our CT-based framework. Agarwal et al. [27] uses the quantization approach on COVID-19 CT analysis but focuses on the segmentation and not classification tasks. The size reduction (36-66% reduction) is significant, but segmentation focus, various measures of evaluation (Dice score vs. accuracy) and cloud-based implementation predetermine the impossibility of direct performance comparison. Yen and Tsao [31] show lightweight CNN optimization with a compressed size 68 percent on medical imaging, but in the case of liver ultrasound classification, not chest CT, so the technology cannot be applied to diagnose thoracic diseases. The suggested integrated knowledge distillation and quantization structure will cover a number of urgent gaps found in the literature aimed at deployment:

- **CT-Specific Optimization**: Unlike existing deployment-focused studies that primarily address X-ray imaging (Termritthikun et al.) or other modalities (Yen & Tsao), our approach specifically targets chest CT classification challenges.
- **Focus on Classification:** Although segmentation-based methods (Agarwal et al.) have their value, binary classification at a glance is one of the most important elements of the clinical deployment, as our framework is explicitly aimed at.
- **Comprehensive Architecture Analysis:** Our evaluation across multiple teacher architectures (DenseNet121, EfficientNetB3, NasNetMobile, etc.) provides broader applicability compared to single-architecture studies.
- **Systematic Integration**: The combined application of knowledge distillation and quantization in a unified pipeline represents a novel contribution not demonstrated in existing deployment-focused literature.

The 91.4% accuracy of the proposed work is a trade-off of moderate level (4-7% accuracy loss) to benefit significantly in terms of deployment. This trade off can be explained by the fact that such high- accurateness methods may demand 3-6x more memory and computational means depending on the complexity of their architecture, which is incompatible with resource-constrained clinical settings. The comparison discloses a crucial issue of the modern medical AI study the crisis of deployment. Models with high performance are not practically applicable to clinics because of computer limitations, and only a limited number of studies that are deployment-oriented can be found, which makes a big gap in the search of a real-world solution to the problem of chest CT classification. The proposed work covers this gap by showing that competitive diagnostic performance (91.4% accuracy) at with extraordinary computational efficiency (75% memory reduction), can be attained, namely when classifying chest CT. This is the first structured system to deployment-optimized deployment-optimal chest CT classification, which offers clinical-grade performance. The state-of-the-art comparison shows that our combined knowledge distillation and quantization framework is at a special place in the literature. More accurate approaches are available, but they do not take into account the feasibility of deployment at all. To fill this serious gap in the research, there are only a few studies on deployment awareness that are based on the specific problem of chest CT classification. The 91.4% accuracy with 618 MB memory footprint of proposed work gives the best trade-off between the diagnostic performance and the ability to deploy chest CT classification. The 75 percent memory loss with competitive accuracy sets a novel standard of deployable medical AI in thoracic imaging, which offers a viable avenue to clinical deployment in a resource-constrained setting. Above all, our multisystem architecture comparison and systematic optimization methodology offers a re-producible system that can be used in other clinical imaging problems, which is part of the long-term objective of transforming medical AI into a viable system, but not just an impressive academic achievement.

## 6. Conclusion and Future Work

The main issue of medical AI implementation is the challenge of balancing between high-performing research models and computationally realistic clinical applications, especially when it comes to chest CT classification, where models such as DenseNet121 demonstrate 100 percent training accuracy but pronounceably reduced validation performance (to overfitting). Our minimalist system of integrated knowledge distillation and post-training quantization is a systematic approach to this problem, which reduces complex teacher models to lightweight, generalisable student networks with clinical grade diagnostic accuracy. The overall analysis of seven state-of-the art architectures shows impressive results: NasNetMobile student scored 93.0% validation accuracy over the teacher 63.5%, EfficientNetB3 student scored 92.0% over the teacher 49.2% and the convergence of training was perfect without overfitting across all the architectures. Post training quantization delivered the same 75% memory reduction (between approximately 2466mb and approximately 618mb) with only a slight degradation on accuracy, with DenseNet121 and Xception quantized models demonstrating slight improvements of 1.4 and 1.1 respectively, reflecting a positive effect of regularisation. The approach effectively resolves the deployment feasibility crisis highlighted by the existing medical AI research, as the models with high accuracy (Ibrahim et al.: 98.05, Malik et al.: 96.67) cannot be used in clinical practice because of computational limitations but do not give deployment statistics. Our framework is the initial systematic deployment-optimized chest CT classification that can preserve the competitive diagnostic performance and provide a viable clinical application because of the significant resources-saving options. This unified methodology forms the initial methodical structure of deployment optimized chest CT classification with the optimal balance between diagnostic precision (91.4) and computational efficiency (618 MB memory footprint) necessary to deploy in any real-life clinical setting deploying mobile CT units, rural health facilities and emergency departments where both the diagnostic performance and resource efficiency are paramount aspects.

## Dataset Availability

The dataset is available at https://www.kaggle.com/datasets/mohamedhanyyy/chest-ctscan-images

## Ethics approval statement

NA

## Patient consent statement

NA

## Permission to reproduce material from other sources

NA

## Clinical trial registration

NA

